# Ventral Tegmental Area as the Core Pathophysiological Hub in Bipolar Disorder: Evidence from Resting-State fMRI

**DOI:** 10.1101/2025.02.08.25321931

**Authors:** Tien-Wen Lee

## Abstract

**Objectives:** Bipolar disorder (BPD) is a major and complex psychiatric condition, clinically characterized by euthymic, depressive, and manic phases. Through the concept of “dialectic neuroscience,” the ventral tegmental area (VTA) has been proposed as a potential key hub in BPD. This study utilized resting-state functional magnetic resonance imaging (rsfMRI) to investigate functional connectivity (FC) changes seeded by the VTA in BPD.

**Methods:** MRI data from 98 participants in a publicly available dataset were analyzed (49 stable BPD patients and 49 healthy controls). Using the dopamine receptor D1 distribution template from JuSpace, the representative voxel of the VTA in the midbrain was identified based on the highest correlation with the D1 template. This voxel served as the seed for FC map construction. The FC maps were Fisher-transformed and compared statistically using an independent two-sample t-test.

**Results:** Significant deviations in BPD were observed in brain regions that were highly consistent with the projections of the VTA, including the ventral and medial prefrontal cortex, orbitofrontal cortex, anterior cingulate cortex, and nucleus accumbens.

**Conclusions:** The FC analysis provided empirical support for the VTA theory in BPD. The role of VTA abnormalities in contributing to the manifestation of opposing polarities within a single disorder was discussed, offering insights into the potential physiological mechanisms and genesis of BPD. The framework of dialectic neuroscience may provide a valuable approach for developing guiding brain theories in psychiatric disorders.

## Introduction

Bipolar disorder (BPD), as its name suggests, is characterized by two opposing phases: mania and depression, and is also known as manic-depressive illness. Brain science research has significantly contributed to identifying consistent patterns of decreased frontal and enhanced limbic neural activities across all three states of BPD (depressive, manic, and euthymic) ^1–3^. Additionally, bipolar mania has been associated with increased baseline metabolism in the striatum, including the ventral striatum (VS) ^3^, while euthymic BPD is also linked to reward sensitivity and negative risk-taking behaviors ^4^. These findings suggest that abnormalities in the reward-related circuitry and basal ganglia (BG) also constitute an enduring neural feature of BPD. However, the underlying disease mechanisms that account for the holistic picture remain elusive. It is evident that neither limbic hyperactivity, including the associated cortico-striato-thalamo-cortical loop, nor a hyperdopaminergic state with enhanced BG activity alone sufficiently explains the occurrence of manic and depressive phases, respectively. Consequently, the pathogenic locus of BPD likely resides outside the limbic region and BG, while still exerting influence over both structures.

Psychiatric disorders are inherently complex, posing substantial challenges in deciphering their fundamental nature. To address this complexity, the author developed the framework of “centralized dialectics” and its application to brain science—termed “dialectic neuroscience.” This approach was previously applied to explore the neuropathology of major depressive disorder (MDD) and has recently received empirical support ^5–7^. Building on this framework, a guiding theory was proposed, identifying the ventral tegmental area (VTA) as a key hub in BPD ^8^. Notably, the neural pattern of hypofrontality and limbic hyperactivity and clinical manifestation of depression are shared by both MDD and BPD ^9^, highlighting a potential transdiagnostic mechanism. Insights gained from deciphering MDD have paved the way for investigating BPD within this theoretical VTA model. This study utilized resting-state fMRI to examine the functional connectivity (FC) patterns centering on VTA in individuals with BPD.

Located near the midline on the floor of the midbrain, the VTA serves as the primary source of dopaminergic cell bodies within the mesocorticolimbic dopamine system. As an inherent dopaminergic hub, the VTA is a plausible candidate for explaining the dopaminergic dysregulation implicated in BPD ^10^. Additionally, the VTA interacts with the ventral striatum (VS) and nucleus accumbens (NAc), which function as relay stations within the limbic-striato-thalamo-cortical pathway ^11^. The rationale for the VTA-centered theory of BPD is briefly outlined below from two perspectives: (1) the projection distribution of the VTA and (2) its physiological properties. Firstly, the VTA has direct connections with the medial prefrontal cortex (mPFC) and the anterior cingulate cortex (ACC), forming the mesocorticolimbic pathway ^12,13^. Moreover, the VTA exhibits strong reciprocal interactions with the VS/NAc, where information from cognitive (PFC), motivational (VTA), contextual (hippocampus), emotional (amygdala), and arousal-related (midline thalamus) sources is adaptively integrated ^14,15^. These VTA-associated pathways robustly modulate neural activity within both the limbic system and the BG, positioning the VTA as a key structure for modeling the opposing poles of BPD. For a more detailed discussion on network interactions in manic and depressive states, see ^5,8^. Secondly, similar to the VS/NAc, the VTA has also been implicated in reward and reinforcement processes ^16,17^. It serves as a central node within a distributed reward-related network, which includes raphe nucleus (RN), lateral habenula, supramammillary nuclei, rostromedial tegmental nucleus, and bed nucleus of the stria terminalis, among others ^17^. Beyond its traditional functions, the VTA is also involved in aversion regulation ^12,18^. Aversive and stressful experiences, such as social threats, foot shock, physical restraint, resource limitation, and suboptimal caregiving environments, have been shown to enhance dopaminergic activity within the mesolimbic system ^12^. In summary, the VTA is engaged in both rewarding and aversive processes, also mirroring the bipolar nature of BPD. Notably, the projection and physiological profiles exhibiting “bipolarity” are features that the neural nodes in the limbic system or BG lack, despite strong evidence linking abnormalities in these regions to BPD.

Since directly localizing the VTA using structural MRI is challenging, the author utilized JuSpace, which provides nuclear imaging templates derived from radiolabeled ligands of various neurotransmitter systems. The foundational paper on JuSpace demonstrated that drug-induced spatial alterations in brain activity correspond to the distribution of specific receptor systems targeted by the respective compounds ^19^. Specifically, receptor density distributions detected by Positron Emission Tomography or Single Photon Emission Computed Tomography have been shown to correlate with neural metric maps obtained from functional MRI (fMRI).

The validity of JuSpace has been established in neurological conditions, such as Parkinson’s disease ^19^, and in the localization of small subcortical nuclei ^20^. Similar to the latter approach, the author utilized a D1 receptor density map to identify the VTA within the midbrain ^21^. The VTA-seeded FC maps of BPD and healthy controls (HC) were then compared to examine potential deviations in the interaction with mPFC, ACC, and NAc ^11–13,17^.

## Materials and Methods

### Subjects, MRI data, and preprocessing

MRI data from 98 participants from the University of California, Los Angeles (UCLA) Consortium for Neuropsychiatric Phenomics LA5c study was obtained ^22^, including 49 BPD patients (with the full dataset being utilized) and 49 HC. At the time of scanning, all BPD patients were in a stabilized condition for at least four weeks. This publicly available dataset can be accessed via OpenNeuro (https://openneuro.org/datasets/ds000030). Whole-brain functional and structural MRI (sMRI) images were acquired using 3.0 Tesla MRI scanners (Siemens Allegra) with a TR of 2s, a voxel size of 3×3×4 mm³, and 152 volumes.

rsfMRI preprocessing was conducted using the Analysis of Functional NeuroImages (AFNI) software package ^23^. The preprocessing pipeline included despiking, slice-time correction, realignment (motion correction), registration of T1 anatomy, transformation to Talairach space (TT_N27+tlrc; both functional and structural MRIs), spatial smoothing, and bandpass filtering at 0.01–0.10 Hz. The first 4 scans were discarded to allow for signal equilibrium. A smoothness kernel of 6 mm was applied to the rsfMRI data. For anatomical segmentation, FreeSurfer was employed to delineate gray and white matter from T1-weighted images and to parcellate the cortical surface into 68 regions of interest (ROIs) using the Desikan-Killiany Atlas. The segmented results were also transformed to align with standard template. This approach facilitated the identification of the midbrain and VTA. To account for motion and physiological noise, 12 movement-related parameters, as well as white matter and ventricular signals, were included as nuisance regressors in the model following established denoising protocols. Additionally, second-order polynomials were applied to model baseline drift.

### Identification of VTA by D1 receptor map

The brainstem ROI delineated by FreeSurfer served as the initial reference for VTA identification. Given that all MRI images were aligned to the Talairach template, it was reasonable to assume that the midbrain lies above the Z coordinate of −30, as illustrated in Figure 1 (subplot A). The time series of each voxel within the midbrain ROI was extracted and correlated with the rest of the brain. A cortical mask was then constructed based on known VTA projection areas, encompassing the ACC, lateral and medial orbitofrontal cortex (lOFC and mOFC), and ventromedial prefrontal cortex (vmPFC) ^12,13^. The correlation between the D1 receptor map and the VTA-FC map was constrained within this mask. The FC map of the voxel showing the highest correlation with the D1 receptor map was considered a surrogate for the VTA. Notably, the BG was excluded to minimize potential confounds from nigrostriatal projections, which could influence the D1 receptor distribution and reduce the sensitivity of the localization strategy. This approach is consistent with existing methodologies utilizing JuSpace, a toolbox designed for cross-modal correlation of MRI-derived measures with nuclear imaging-based estimates of various neurotransmitter systems ^19,20^.

**Figure 1.**
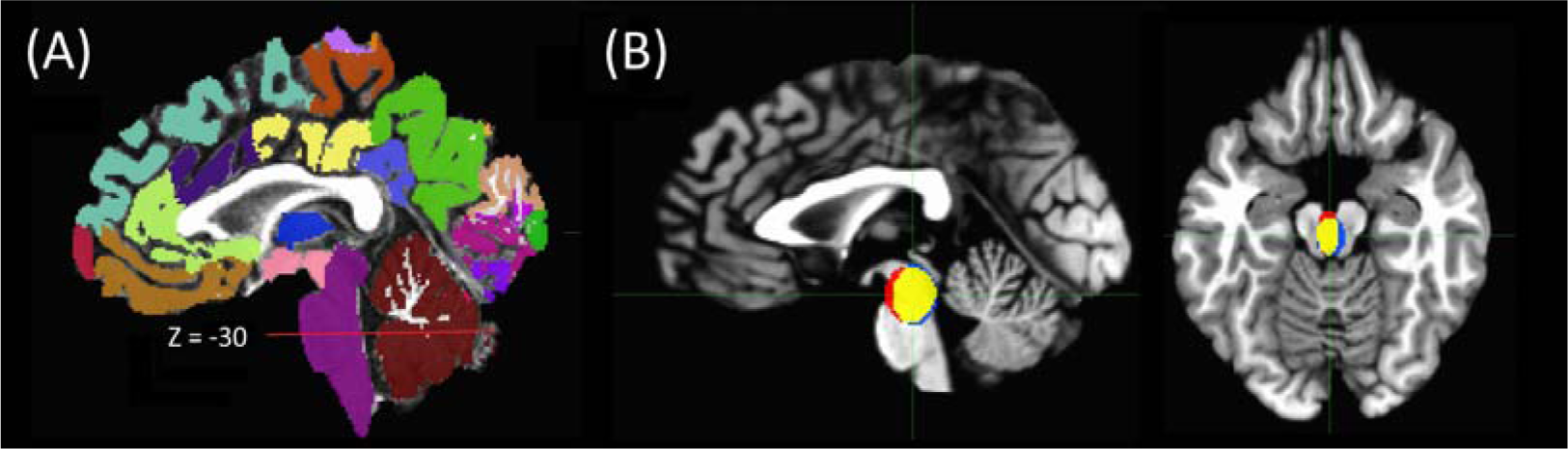
(A) Segmented structural MRI image from a selected case for demonstration. The purple ROI indicates the brainstem, and the red line marks the Z coordinate at −30, with the midbrain structure located above the line. (B) Target VTA voxel positions represented as ellipsoids with dispersion of 1 SD along the three axes for the BPD and HC groups, superimposed on a standard Talairach template. Blue: BPD, Red: HC, Yellow: overlapped. Sagittal and axial views are centered at [0.2, 23.1, −15.1], the global mean coordinate of the target VTA voxels across all participants. This distribution aligns with the elongated structure of the VTA in the midbrain.

To assess whether there is a significant difference in the distribution of VTA coordinates between the BPD and HC groups, Hotelling’s T² test was employed, which is a multivariate generalization of the t-test used for comparing the means of two groups with more than one variable (in this case, the 3D coordinates: x, y, and z). The test involved the calculation of respective sample mean and covariance matrix. The pooled covariance matrix of the two groups was computed. Hotelling’s T² statistic was then calculated using the formula, which incorporates the difference between the group means, the pooled covariance matrix, and the sample sizes of both groups. This statistic quantifies the multivariate distance between the two group means, adjusted for the variance and covariance structure. A P-value less than 0.05 indicates that the two groups’ distributions of VTA coordinates differ significantly.

### FC map construction and statistical comparison

The voxel values of the VTA-centered FC maps were transformed into Fisher’s z-scores prior to group-level statistical comparisons (where a Fisher’s z-score of zero corresponds to a correlation coefficient of zero). A two-sample independent t-test was conducted to identify significant brain regions with scores showing between-group differences using the AFNI module 3dttest++ ^23^. The statistical threshold for the t-test was set at a corrected P-value < 0.05 (non-parametric alpha permutation) with an extent threshold > 30 voxels. Given that the BPD patients were not in active disease states and hence that the between-group differences were expected to be less robust, a more lenient uncorrected P-value < 0.005 was also reported.

## Results

The mean ages of the BPD and HC groups were 35.3 years (SD = 9.0) and 35.3 years (SD = 8.4), respectively, with no significant between-group difference (P = 0.991, t = 0.012, df = 97). The gender distributions for the BPD and HC groups were 28/49 and 26/49, respectively, with no significant distribution difference (Chi-Square statistic = 0.165, P = 0.685). As for other protocol details, please refer to the data source ^22^.

The average coordinates of the representative VTA for the BPD and HC groups were [−0.34, 23.8, −15.3] and [0.75, 22.3, −14.8], respectively, following the RAI convention, on the Talairach brain template (TT_N27+tlrc provided by AFNI), as shown in Figure 1 (subplot B). The square roots of the eigenvalues of the respective covariance matrices, which represent the spatial dispersion, are [6.2, 7.4, 10.3] and [4.6, 8.8, 9.6]. It is noteworthy that one major contribution to the dispersion comes from the imperfect (which is a fact) alignment of fMRI to sMRI and to the template. The distribution of the identified VTA loci did not show any group differences, with the mean coordinate difference approaching zero, with Hotelling T² statistic: 2.0806 and P-value 0.563.

The VTA-centered FC maps demonstrated significant differences in the vmPFC (including the frontal pole), bilateral OFC (including pars orbitalis), anterior cingulate cortex (ACC), and NAc, which were in good concordance with the major cortical and subcortical projections of the VTA. The results are illustrated in Figures 2 and 3 and summarized in Table 1.

**Figure 2.**
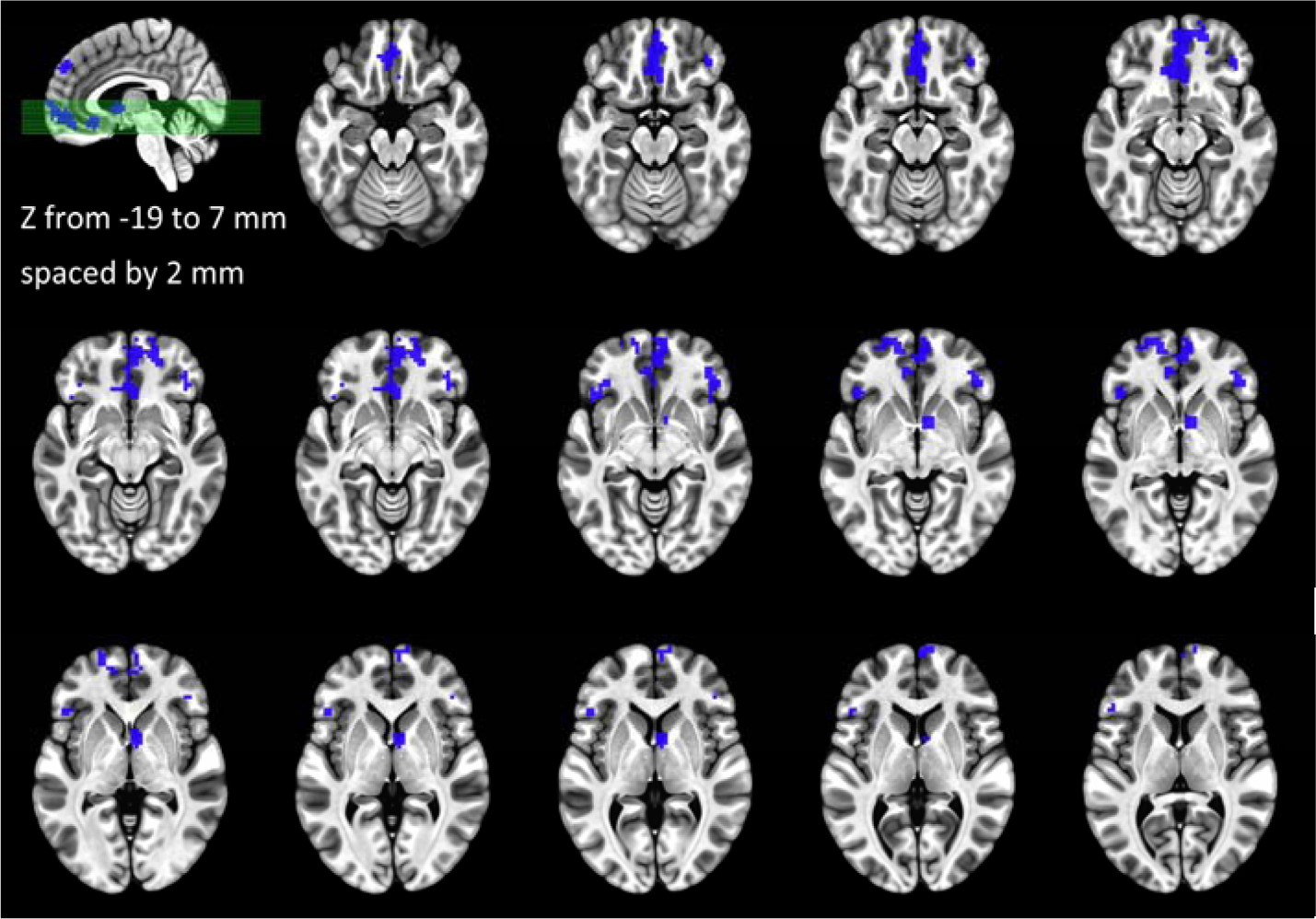
Mosaic axial views depicting the abnormal clusters of the VTA-seeded FC map. Blue indicates decreased functional connectivity in the BPD.

**Figure 3.**
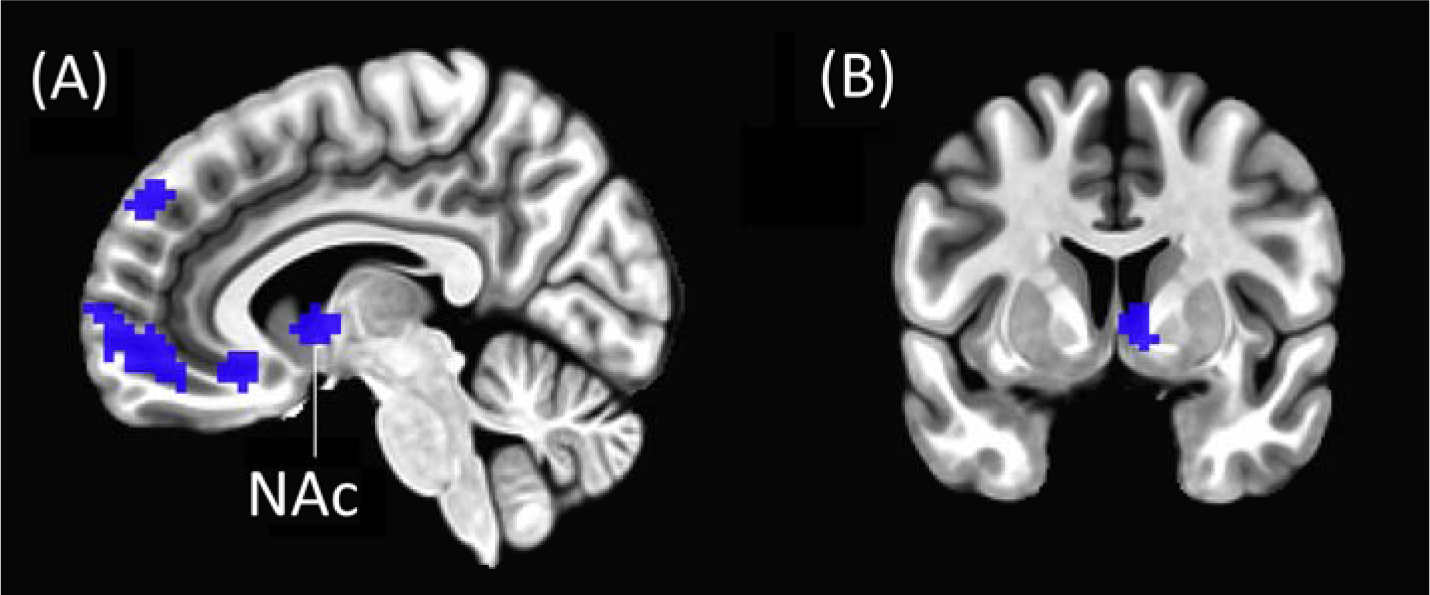
Sagittal and coronal views for better illustration of the abnormal clusters in the medial PFC (subplot A) and NAc (subplots A & B) in BPD. Please refer to Table 1 for the detailed information of the coordinates and statistics.

**Table 1.**
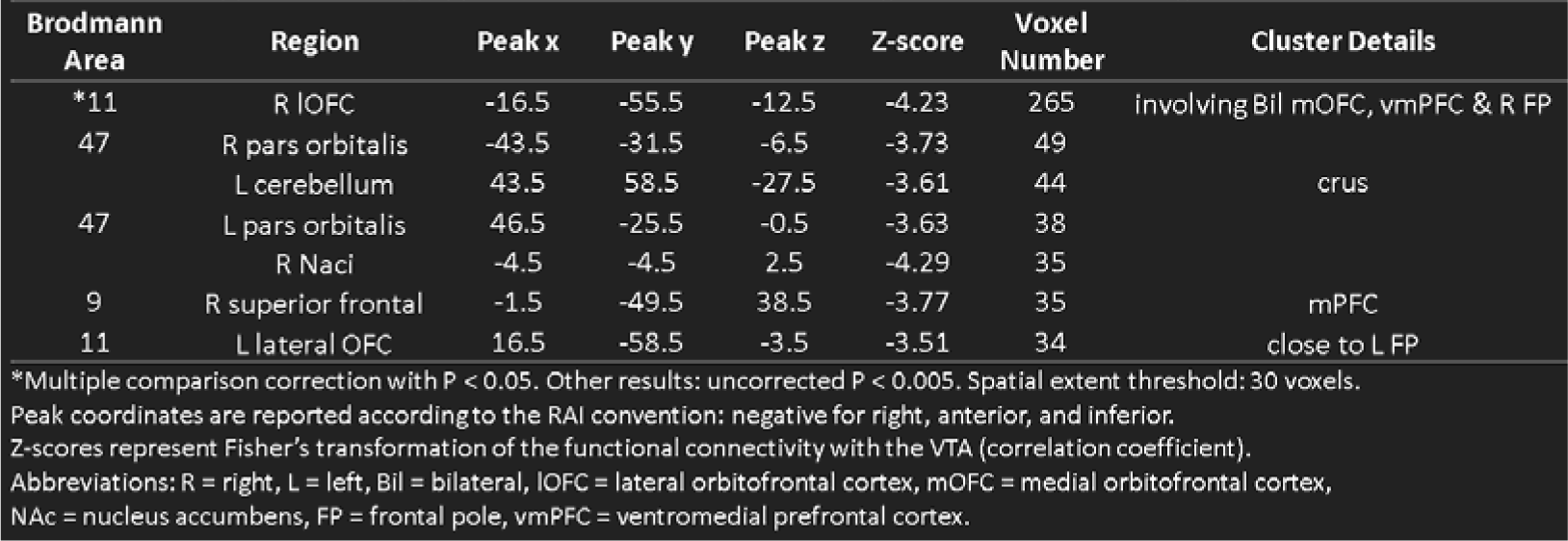
Brain regions showing significant functional connectivity changes in BPD compared to HC.

## Discussion

While substantial evidence supports dopaminergic hyperactivity in mania, the presence of intervening depressive and euthymic phases in BPD complicates the identification of the core abnormality’s precise location. Hyperdopaminergism alone seems insufficient to explain the observed decrease in frontal activity and increase in limbic activity across the three phases. Previous research has made commendable efforts to investigate BPD from various perspectives, including emotion regulation and behavioral activation systems ^24^. However, a comprehensive explanation of the disease mechanism and pathogenesis has been lacking. This research provides empirical evidence supporting the theory that the VTA serves as a central hub in the pathogenesis of BPD ^8^. Verification of this hypothesis has been hindered by two major factors. First, the challenge of localizing the VTA using in vivo neuroimaging tools. Second, the complexity of depression pathogenesis. Recent advancements integrating functional and neurochemical imaging techniques, along with a deeper understanding of MDD pathogenesis from a brain science perspective (the latter to be elaborated later), have addressed these obstacles ^6,19^. With the D1 receptor template provided by JuSpace, a surrogate VTA position was elucidated, and the FC map with VTA as a seed was constructed and compared. Interestingly, the brain regions showing significant deviations in BPD were highly concordant with the projections of VTA, including mPFC, vmPFC, OFC, ACC, and NAc. These findings persuasively support the role of VTA in BPD. In addition, samples from stabilized BPD patients suggest that abnormal VTA activity could be an enduring feature of the disorder. However, among the diverse neuroimaging findings of BPD, why is the VTA qualified to be the key hub (a proposal derived from the efforts of dialectic neuroscience ^7,8^)? This question is addressed through an exploration of several key points outlined below.

### Summary of neural abnormalities in BPD

A wide range of neuropsychological domains are affected in patients with BPD ^25–27^. Some deficits, such as impairments in sustained attention, inhibitory control, reward sensitivity, verbal memory, and executive function, may persist even in remitted BPD or BPD with residual symptoms. The persistence of these neuropsychological deficits during euthymia aligns with genetic research suggesting a strong biological basis for BPD ^28^. This broader perspective reinforces the view that BPD has a substantial genetic component, with neuropsychological impairments persisting even in symptom-free periods.

Despite diversities in neuroimaging findings, three major categories of neural abnormalities have been observed in BPD: reduced frontal activity/responsivity, along with heightened activity/responsivity in the limbic system and BG. For example, during the execution of a working memory task, all three BPD phases exhibited a consistent pattern of reduced neural activity in dorsal (cortical) regions and increased activity in ventral (limbic) regions ^1,2^. The findings of hypofrontality and limbic hyperactivity have been replicated across a wide range of cognitive and affective experimental paradigms ^29–31^.

Reward abnormalities in BPD are of particular interest. It has been proposed that a competition exists between the desire for an immediate reward (reflexive or instant gratification) and the delayed satisfaction of a long-term goal (reflective reward or temporary reward suppression). In bipolar mania, both reflexive and reflective reward processes appear to be impaired, with a stronger inclination toward the former and reduced proficiency in the latter. Motivational and reward-based tasks in BPD have been associated with heightened neural responses in the BG (BG), particularly VS ^29,31,32^. Even in the euthymic phase, BPD patients exhibit reward sensitivity and negative risk-taking behaviors ^4^, suggesting that dysfunctions in reward-related circuitry and the BG are enduring features of the disorder. Notably, the BG also show an exaggerated response to affective stimuli ^29,33^, a pattern that persists across all three BPD phases ^34^. From clinical perspective, up to 50 percent of BPD patients may experience psychotic symptoms, and dopamine antagonists are effective at relieving psychosis ^35^. These findings align with the concept of hyperdopaminergism and BG dysfunction in BPD.

### VTA theory to account for the depressive phase in BPD

As summarized above, the primary neural dysfunctions in BPD involve the frontal and limbic regions, as well as the BG/ventral striatum (BG/VS), which are central to the motivational dopaminergic circuitry. Notably, these three categories of abnormalities appear to be distinct and independent, with no clear explanatory link between them. This suggests that the core pathology of BPD likely extends beyond these systems. With the recent understanding that depression arises from a compartment-level network imbalance between the frontoparietal and limbic systems, the findings of hypofrontality and limbic hyperactivity can be integrated into a unified framework ^5,7^. Specifically, a reciprocal suppressive mechanism (RSM) between the frontoparietal and limbic compartments enables dynamic switching between external engagement and internal processing. A disruption of this balance drives the system into an attractor state, manifesting as depression—characterized by limbic hyperactivity and hypofrontality. The cortico-limbic RSM, together with several well-documented network mechanisms, may account for a substantial portion of the understanding of MDD and secondary depression (e.g., post-stroke depression) ^5^. How, then, does VTA theory account for the occurrence of depression in BPD?

Limbic hyperactivity in the euthymic state may stem from both enhanced VTA-limbic interactions and the limbic-VS-thalamo-cortical pathway, given the strong connectivity between the VTA and the VS/NAc. Through the reciprocal suppressive mechanism (RSM), this hyperactivity leads to hypofrontality. When the cortico-limbic imbalance reaches a critical threshold, BPD manifests as depression. Clinically and neurologically, depression in BPD is nearly indistinguishable from MDD due to their shared neuropathological mechanisms. However, from a therapeutic standpoint, strong evidence suggests that depression in BPD and MDD differs. Antidepressants are a primary treatment for MDD; however, their efficacy in bipolar depression is often limited when used as monotherapy and carries the risk of inducing a switch from depression to mania. Instead, several classes of medications have been shown to be effective in treating BPD, including lithium and anticonvulsants, collectively referred to as mood stabilizers. Lithium, one of the oldest treatments for BPD, has an incompletely understood mechanism of action. Proposed mechanisms include inhibition of inositol monophosphatase, modulation of G-proteins, regulation of signal transduction cascades, interaction with nitric oxide signaling, mitochondrial protection from oxidative stress, competition with magnesium to reduce glutamatergic neurotransmission, and promotion of neuroprotective protein release ^36^. Additionally, several anticonvulsants used for epilepsy have demonstrated therapeutic efficacy in BPD. Their mechanisms may involve interference with voltage-sensitive sodium channels, enhancement of GABAergic inhibition, and modulation of signal transduction pathways. Despite differences in their specific mechanisms, mood stabilizers share a common function: stabilizing neuronal membranes and reducing neuronal excitability ^37^, which aligns with the overactive VTA theory. Furthermore, antipsychotics may influence VTA firing through the NAc-to-VTA feedback projection and the dopaminergic autoreceptors on VTA neurons ^38^. The efficacy of antipsychotics in bipolar depression supports the role of dopaminergic dysfunction in BPD, reinforcing the hypothesis that an overactive VTA underlies its pathology. Correcting the overactive VTA and the imbalance between the frontoparietal and limbic compartments may be achieved through a treatment approach combining mood stabilizers and antidepressants. The VTA theory thus offers a compelling framework for understanding the differential pharmacological responses to depression in MDD and BPD.

### VTA theory to account for the manic phase in BPD

The BG are among the primary regions implicated in BPD, particularly the VS in mania. Since the VTA powerfully modulates the activity of the BG ^15,39^, the VTA theory is also ideal to account for the observed BG abnormalities in patients with BPD. It has long been recognized that mania/hypomania is a dysregulated state characterized by heightened sensitivity to reward-related stimuli, increased goal-directed activities, and excessive pleasure-seeking behaviors ^40,41^. In line with clinical and psychological assessments, neuroimaging research has revealed enhanced reward-related neural responses in manic/hypomanic patients or individuals prone to mania ^10,32^. The cortico-striato-thalamo-cortical circuit forms the core structure for the functionally segregated parallel networks of the BG, with cortical areas including the motor, PFC, oculomotor, and limbic regions ^42^. The VS acts as a relay station within the limbic-striato-thalamo-cortical pathway and has reciprocal interactions with the VTA. Unlike other parallel pathways of the BG, the connections of the VS are more intricate, extending to deep nuclei such as the VTA, hypothalamus, hippocampus, amygdala, and RN. Furthermore, the VS/NAc can modulate dopaminergic signaling in other striatal regions by projecting to the VTA and substantia nigra through the spiraling striato-nigro-striatal circuitry ^39^. This organization enables interactions between the parallel cortico-striato-thalamo-cortical circuits, facilitated by projections from the VS-VTA pathway, which may also influence neural activity in other PFC regions beyond the vmPFC. Concordantly, increased PFC activity in bipolar mania compared to the euthymic state has been observed ^1^. In addition, the heightened activity in the BG and PFC during mania may mitigate limbic hyperactivity via RSM. This hypothesis aligns with neuroimaging findings showing reduced amygdala responses to negative emotional stimuli in mania relative to the euthymic state ^34^. Thus, in bipolar mania, the cortico-limbic balance remains intact due to enhanced BG/PFC activity. Overall, the hyperdopaminergic state of the BG emerges as a key neural signature, driven by multiple sources: direct VTA inputs, frontal inputs via the fronto-striatal pathway, limbic inputs via the VS-VTA pathway, and less RSM from the limbic system.

Under the assumption of an overactive VTA, why might depression occur independently of mania? In depression, pronounced limbic hyperactivity may attenuate frontal activity via the RSM, leading to reduced frontal input to the BG, which in turn may counterbalance the increased limbic input. Additionally, excessive limbic activation could trigger compensatory feedback from the vmPFC and subgenual ACC to the RN, dampening the reward system ^9^, including the VTA. This explains why BPD can manifest as pure depression (nearly indistinguishable from MDD), where the primary neural signature is unbalanced hypofrontality and limbic overactivity, without excessive BG activation. Accordingly, the presentation of depression or mania is shaped by the interaction and competition between large-scale networks or by the dynamic regulation of VTA neuronal subpopulations (see the section *Physiological mechanisms related to the genesis of BPD* for details). Following this reasoning, mixed-type BPD represents a state where cortico-limbic imbalance coexists with hyperdopaminergic BG activity. The hyperactive BG may arise from a particularly strong or efficient VTA-VS drive (which remains uninhibited by hypofrontality and negative feedback from the RN), together with mPFC input via the fronto-striatal circuit. Based on this framework, the VTA model of BPD can accommodate opposite affective polarities within a single disorder.

Increased limbic activity in BPD can be explained by the VTA to limbic projection outlined in the previous section. The heightened VS activity driven by the VTA may further amplify limbic cortical activity via the cortico-striato-thalamo-cortical circuit, providing an additional source of limbic hyperactivity. These pathways of limbic enhancement may account for the cortico-limbic imbalance being biased toward the ventral compartment across all three disease phases and for the greater prevalence of depressive episodes compared to manic episodes in BPD.

### Physiological mechanisms related to the genesis of BPD: positive feedback circuits and overactive VTA

About 60–65% of the VTA neurons are dopaminergic, while the remaining 35% are GABAergic ^16^. The dopaminergic neurons in the VTA are phasically active and exhibit repetitive burst-like firing, which is closely related to their physiological function. Interestingly, the neural circuits surrounding the VTA contain multiple positive feedback mechanisms, which may contribute to the persistence of neural abnormalities in BPD. For instance, projections from the NAc to the VTA primarily target GABAergic neurons, resulting in a net disinhibition of VTA dopaminergic firing (i.e., inhibition of inhibition via GABAergic neurotransmission) ^12^. Similarly, afferents from the mPFC and limbic regions to the VTA can enhance VTA dopaminergic activity through glutamatergic projections from the mesolimbic system to the VTA or VTA-VS ^12,43^. Furthermore, the cortico-striato-thalamo-cortical loop, a fundamental BG pathway, may also function as a self-reinforcing circuit, further amplifying these effects (note: the mechanism is complex, with one possibility involving the differential distribution of D1 and D2 receptors in the mPFC and striatum ^44^). The advantage of these positive feedback circuits lies in their ability to sustain prolonged goal-directed and motivation-driven behaviors, mediated by the NAc (motivation), caudate (planning), and putamen (execution) ^11^. This autofeedback design, with the VTA as a key ignitor, is adaptive and essential for survival and is finely regulated by a complex array of neural inhibitory processes. However, a potentially disastrous consequence is “overheating.” In the case of bipolar mania, over-activation of the BG may be driven by the VTA (and/or VTA-VS). Additionally, iatrogenic mania can emerge when autofeedback circuits are “hacked” by medical interventions, such as deep brain stimulation or pharmacological treatments like levodopa. Overheating of VTA-BG structures may give rise to hallmark symptoms of mania, such as hypertalkativity, racing thoughts, heightened goal-directed activities, and excessive pleasure-seeking behaviors. This dysregulation is often accompanied by an expanded sense of self, potentially manifesting as inflated self-esteem or grandiosity. Notably, while autofeedback mechanisms may contribute significantly to the persistence and expression of BPD, the core pathology of BPD likely lies beyond these adaptive circuits and instead stems from an overactive VTA, further discussed below.

“Bipolarity” is a defining feature of BPD, which not only manifests in network activities, as discussed above, but also exists within the VTA itself. For example, susceptibility to depression has been linked to opposing trends in phasic firing between mPFC-projecting and NAc-projecting VTA neurons ^45^. Beyond reward signaling, stress-related neuroendocrine molecules—such as cortisol, corticotropin-releasing factor, and dynorphin—also exert significant effects on VTA neurodynamics, which can occur within several hours ^12^. Although an overactive VTA appears to be a promising candidate as a core pathological locus of BPD, its underlying molecular and physiological mechanisms remain unknown. It is noteworthy that manic or depressive episodes can occur spontaneously and are not necessarily preceded by stress. The “kindling” phenomenon of BPD—characterized by progressively shortened disease-free intervals—suggests that the underlying abnormalities accumulate with age or with the number of episodes. Therefore, the neural aberrations in the VTA of BPD follow a progressive or cumulative trajectory.

Dopaminergic neurons in the VTA fire spontaneously in a pacemaker-like manner, a process supported by several mechanisms, including the large h-current (I[) mediated by hyperpolarization-activated cyclic nucleotide-gated channels ^46^. The molecular basis of the sensitization or kindling-like phenomenon could be linked to glutamatergic mechanisms and the plasticity of N-methyl-D-aspartate (NMDA) receptors ^47^. Similar to epilepsy, pathology may accumulate to a critical threshold, at which point large-scale network stability is disrupted, leading to the emergence of depression or mania. Below this threshold, VTA abnormalities do not cause overt impairment in BPD patients and can only be detected through neurophysiological or psychological assessments. From an evolutionary perspective, autofeedback mechanisms aiming for active working mode may align with circadian rhythms. It is thus also reasonable to assume that the proposed gradual increase in VTA oscillatory fluctuations could be linked to chronobiological dysregulation. Chronobiology encompasses a range of rhythms, including daily (diurnal), tidal, weekly, seasonal, and annual cycles. The physiology and behavior of an organism can undergo significant changes depending on the phase of its biological rhythm. Notably, a substantial proportion of BPD patients exhibit seasonal patterns. Recent research highlights that nearly all cells and tissues in the body function as biological clocks, expressing the Clock gene (circadian locomotor output cycles kaput), not just the suprachiasmatic nucleus ^48^. The molecular foundation of circadian rhythms is built upon Clock proteins (both activators and repressors), as well as associated kinases and phosphorylases. Interestingly, human studies have suggested an association between Clock genes and BPD ^49^. Moreover, animal research demonstrates that the ClockΔ19 mutant mouse model of mania exhibits increased activity in VTA dopaminergic neurons, further implicating circadian mechanisms in the pathophysiology of BPD ^37^. Additionally, the VTA is a heterogeneous structure, and its dopaminergic neurons can be organized into clusters based on differential projection profiles ^50^. Whether abnormal interactions between these sub-groups of VTA neurons also contribute to the gradual enhancement in VTA warrants further animal and electrophysiological studies to elucidate.

In summary, similar to MDD, BPD also reflects a “bug” in brain organization:

MDD is the price paid for expanded and integrated mental capabilities that compete with each other (i.e., cortico-limbic interactions via the RSM) ^5,6^, and BPD is the potential consequence of having a neuroanatomical design that aims to support sustained actions and explorations (for minutes or even hours). The neural pathways for the occurrence of depression and mania in BPD are tentatively summarized in Figure 4. Gradually enhanced neurotransmission or firing of the VTA is the very likely neural pathology of BPD. Future studies of BPD may place more emphasis on this midbrain structure that has long been ignored in its brain science research.

**Figure 4.**
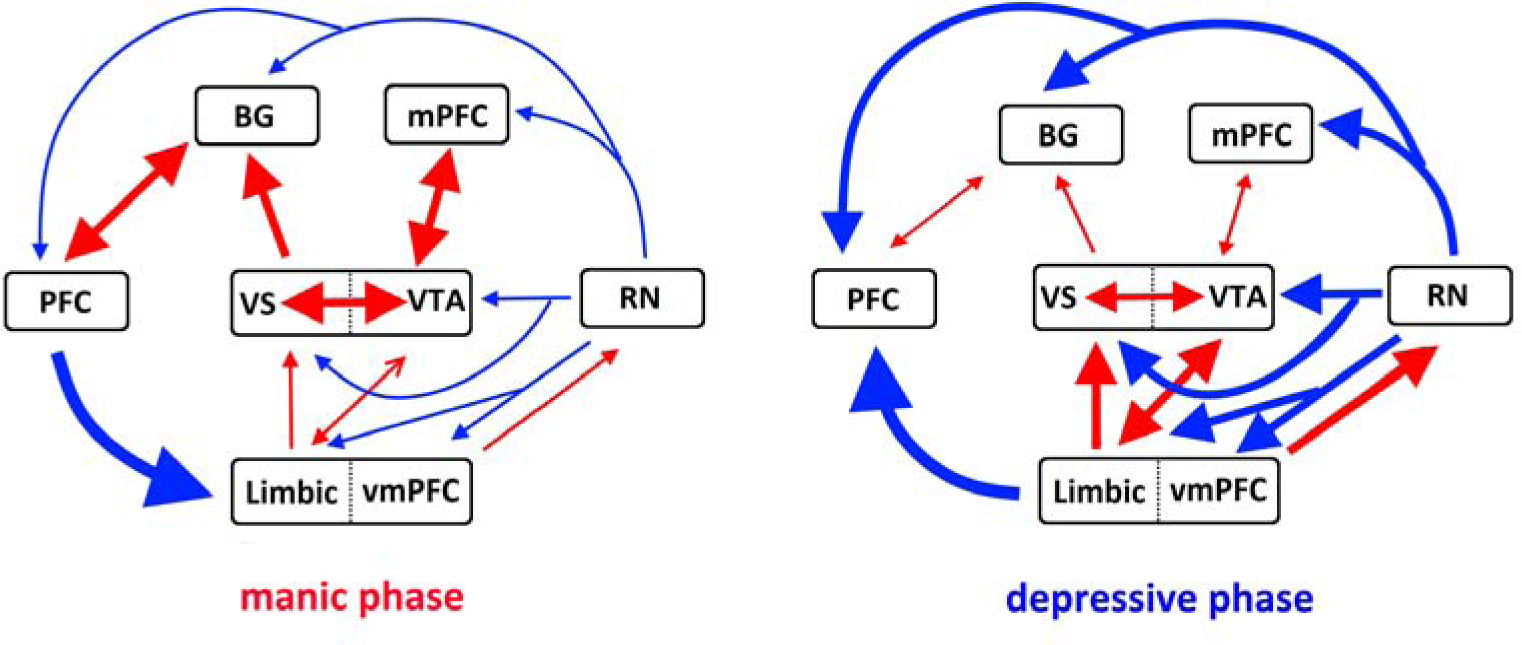
Tentative VTA-related pathways leading to mania and depression are summarized in the left and right subplots, respectively. Reproduced from the author’s own work (Figure 3 in ^8^), permitted under open-access terms. Abbreviations: BG = basal ganglia, PFC = prefrontal cortex, mPFC = medial PFC, vmPFC = ventromedial PFC, VS = ventral striatum, VTA = ventral tegmental area, RN = raphe nucleus. Red: activation. Blue: suppression. Thickness of the lines conceptually represent the strength of interaction.

### Conclusion

This study provides empirical evidence supporting the theory formulating VTA as a key hub in the pathogenesis of BPD. The superiority of the VTA model is supported by its unique neuroanatomical properties, namely its strong innervation of both the BG and the limbic system. Furthermore, the VTA plays a crucial role in both reward and aversive conditions. These make an “over-driving VTA” a compelling candidate for the psychopathological mechanism capable of accounting for both manic and depressive states. The identification of the VTA relied on the state-of-the-art D1 receptor template from JuSpace, along with functional connectivity analyses of rsfMRI. According to the VTA theory, the bipolar nature of BPD may originate from within the VTA itself and from its large-scale network interactions. Whether depression or mania is presented is contingent on the network dynamics, which may, in part, be pre-configured within the VTA—specifically, the overall electrophysiological activity of its distinct neuronal subpopulations. From theory formulation to empirical verification demonstrated in MDD and BPD, “dialectic neuroscience”, in contrast to the dominant psychological or computational approach, could provide a valuable framework for developing guiding brain theories for psychiatric disorders.

## Authors Contributions

This report is authored by a single individual.

## Data Availability

All data produced are available online at https://openneuro.org/datasets/ds000030

## Acknowledgments

I would like to express my gratitude to the University of California, Los Angeles (UCLA) Consortium for Neuropsychiatric Phenomics LA5c study for generously sharing the MRI data.

## Financial support

N/A.

## Statements and Declarations

No conflicts of interest to declare.

## Compliance with ethical standards

This research analyzed the databank from a publicly released dataset. The author carried out no animal or human studies for this article.

## Notes

### Competing Interest Statement

The authors have declared no competing interest.

### Funding Statement

This study did not receive any funding

### Author Declarations

The study used ONLY openly available human data that were originally located at: https://openneuro.org/datasets/ds000030

